# SHAPE-AI: Development and Expert Validation of a Survey for Human–AI Performance Evaluation in Healthcare

**DOI:** 10.64898/2026.01.18.26344350

**Authors:** Mark V Mai, Mustafa Ozkaynak, Jenna L. Marquard, Richard J. Simonson, Richard J. Holden, Hanna J. Barton, Ken Catchpole, Melinda Jamil, Sarah Brauer, Swaminathan Kandaswamy

## Abstract

**Objective:** To develop and content-validate a brief, expert-informed **<u>S</u>**urvey for **<u>H</u>**uman–**<u>A</u>**I **<u>P</u>**erformance **<u>E</u>**valuation (SHAPE-AI) for near-real-time assessment of how clinical AI affects human performance.

**Background:** AI-enabled clinical decision support can improve outcomes only when aligned with clinician workflows, and cognitive demands. Existing evaluations measure technical performance and adoption , providing limited assessment of how AI shapes human performance. There is a lack of concise, operationally feasible instruments to measure AI impact on these human factors outcomes in clinical settings.

**Method:** We used a construct-driven, multi-stage development process. A literature review and prior qualitative work with users of a deployed sepsis prediction tool identified core human performance constructs. Preliminary items were created and iteratively refined through two expert panels. Six clinical informatics experts evaluated representativeness and clarity using content validity indices (CVI). Seven human factors experts then refined constructs, item wording, and response formats through ratings and focus groups, emphasizing discriminant validity, cognitive bias mitigation, and feasibility for deployment within 24 hours of AI use.

**Results:** A concise 10-item instrument was created, comprising perceived impact, interpretability, agreement with the AI’s findings, agreement with the AI’s recommendations, trust, workload, provider–patient and provider–team relationships, unexpected outcomes.

**Conclusion:** The SHAPE-AI instrument is a theoretically grounded, operationally feasible tool to monitor human performance as relates to AI use.

**Application:** Health care organizations can deploy SHAPE-AI as a rapid, standardized probe to detect workflow misalignment, mis-calibrated reliance, communication disruptions, and unintended consequences of AI, informing safer design, implementation, and optimization of clinical AI tools.

*Précis:* SHAPE-AI is a brief, expert-validated survey designed to capture clinicians’ near-real-time perceptions of how AI-enabled decision support affects human performance such as their situational awareness, decision-making, workload, and trust. SHAPE-AI offers health systems a practical, standardized way to monitor and understand the impact of AI on human performance.

## INTRODUCTION

Artificial intelligence (AI)–enabled tools are increasingly deployed in healthcare to improve clinical outcomes.^1^ However, good outcomes are only possible when these tools fit clinician workflows and the broader sociotechnical context in which they are embedded. A model may be mathematically robust yet still fail clinically if its outputs are hard to interpret, appear at the wrong time, or erode user trust.^2,3^ In this sense, technical accuracy is necessary but not sufficient for a safe, high quality care delivery.

Despite this reality, the implementation of clinical AI is currently hindered by three systemic failures: an over-emphasis on technical accuracy, a neglect of workflow integration, and a failure to account for the complexities of human cognition

First, technical accuracy describes how well the model predicts or classifies clinical events, often measured by discrimination and calibration.^3^ These metrics are the focus of most AI evaluations. They tell us how the model performs with data, but not whether clinicians will see, understand, or use its outputs in real time.

Second, a neglect of workflow integration often renders even accurate models unhelpful; an alert that fires too often, appears at a busy moment, or is buried in a crowded interface can disrupt situational awareness rather than enhance it .^4–6^ The same vendor sepsis prediction model has produced very different outcomes in different health systems: one reported increased alert fatigue,^4^ while another reported reduced mortality.^7^ These contrasting results highlight how local workflows, organizational context, and implementation decisions shape whether AI tools are ultimately beneficial.

Finally, human cognition refers to how clinicians perceive, interpret, and act on information from AI tools. AI does not operate in isolation; it becomes part of a joint human–AI system in which clinicians must combine model outputs with their own expertise, patient preferences, and resource constraints.

### Established Human Factors Constructs

Human factors research provides the necessary constructs to explain why AI succeeds or fails at the point of care. One key construct is situation awareness - what information clinicians notice, how they make sense of it, and how they anticipate what will happen next.^8^ An AI tool might improve situation awareness by highlighting a subtle risk that would otherwise be missed, or it might degrade it by distracting attention from more important cues.

Another key construct is trust, the extent to which clinicians are willing to rely on the AI tool when making decisions.^9,10^ If clinicians under-trust the tool, they may ignore useful recommendations. If they over-trust-, they may follow incorrect advice without sufficient scrutiny (over-reliance or automation bias).^9–11^ Communicating limitations and uncertainty about the tool can help calibrate trust and reduce automation bias, supporting more balanced decision making. ^12^

A related construct is interpretability, which we use here to mean how understandable and usable the AI output is to the clinician in context.^13^ A technically “explainable” model is not necessarily interpretable at the bedside. If clinicians cannot see why a patient is being flagged, they may either disregard the alert or accept it uncritically. Another important idea is human–AI agreement: the extent to which clinicians’ judgments align with the tool’s findings and recommendations, and when they reasonably diverge.

Workload and cognitive burden are also critical constructs. AI tools are often promoted as ways to reduce burden, but they can also add work by generating more alerts, requiring extra checks, or prompting new documentation tasks. ^6,14–17^ An accurate alert that appears at a high stress-moment can increase cognitive load and distraction. Over time, this can contribute to alert fatigue and burnout.

AI tools can further affect team communication and relationships, how AI reshapes interactions among clinicians, patients, and the technology itself. ^14^ An AI generated risk score might prompt more timely huddles, or it might replace direct communication with a silent alert in the electronic health record. These changes influence not only efficiency and coordination, but also clinicians’ perceived safety of using the tool in -real-world-practice.

### The Need for Real-Time Measurement

Despite the importance of these human and workflow factors, most evaluations of clinical AI still rely on technical metrics and coarse adoption outcomes, such as alert adherence or override rates.^3^ These measures show whether clinicians followed an alert, but not *why* they did so, or how the AI tool affected their situation awareness, decision-making, workload, or team interactions. In other words, we lack practical ways to measure the “missing link” between model performance and clinical outcomes.

Human factors principles are crucial for understanding not only how clinicians use AI when it works well, but also how they detect, interpret, and recover from AI--related-errors.

Clinicians are often the first to sense when an AI tool is misaligned with real-world practice. By evaluating clinicians’ near-real-time experience of AI at the point of care, health systems can detect latent failures and integration problems before they lead to harm. In this way, clinicians can act as “canaries in the coal mine” for post-deployment performance drops, workflow mismatches, and potential bias in AI tools. There is also a growing risk that differences between AI recommendations and clinicians’ final decisions will be interpreted simply as noncompliance, rather than as appropriate clinical judgment in the face of imperfect tools. Without a clear view of human–AI interaction, clinicians may be unfairly blamed for overriding AI in situations where the model’s recommendation is poorly timed, poorly calibrated, or misaligned with patient needs.

Despite recognition of the need to measure human performance in relation to AI systems,^18^ there remains a paucity of validated, operationally feasible instruments designed to capture these human factors constructs in near-real---time. Existing measures tend to focus on general attitudes toward technology or require lengthy administration that is impractical in busy clinical settings.

### Study Objective

The objective of this study was therefore to develop and content-validate a brief, expert--informed survey instrument to assess the near--real--time impact of AI on human performance in clinical settings, termed SHAPE---AI (**S**urvey for **H**uman-**A**I **P**erformance **E**valuation). Prior work by our group using a deployed sepsis prediction alert identified several key human performance constructs, including changes in situational awareness, diagnostic reasoning, treatment planning, team communication, and response behavior.^19^ Building on that foundation, SHAPE-AI is designed as a concise, generalizable measurement instrument that can be deployed shortly after AI use across different clinical AI and clinical decision support tools. It captures clinicians’ perceptions of impact, agreement with AI findings and recommendations, interpretability, trust, workload, sociotechnical relationships, and unintended consequences. Such an instrument can support rapid---cycle evaluation, guide implementation adaptations, and provide a standardized lens on human–AI teaming that complements technical performance metrics and outcome data.

By utilizing a construct-driven process that integrated prior empirical work, literature review, and sequential input from clinical informatics and human factors experts, we sought to operationalize core domains of human–AI interaction. Our goal was to develop a tool that is both practical for routine deployment and conceptually aligned with contemporary human factors theory, enabling health systems to systematically monitor how AI systems shape clinician decision-making, workload, team dynamics, and perceived safety.

## METHODS

### Developing measurement constructs

Following the survey scale design process described by AMEE (An International Association for Medical Education) Guide No. 87, ^20^ we first conducted a literature review to identify foundational measurement constructs.^18^ As described in our previous published work,^21^ we conducted semi-structured interviews with users of a live sepsis AI system to elucidate how they interacted with the AI tool and understand the operationalization of these human performance constructs in the context of acute care settings. To minimize recall bias, we identified and interviewed providers and nurses within 72 hours of them seeing the AI sepsis tool in production. After synthesizing these qualitative learnings, we developed preliminary survey items.

### Validation and refinement of preliminary survey items

We used the content validity index (CVI) to evaluate how well the items represented each human performance construct being measured. In line with psychometric standards, a CVI of >=0.8 was established as the threshold for item retention and refinement.^22^ We conducted sequential expert validation of the survey instrument with two distinct expert panels to ensure the tool achieved both clinical relevance and human factors rigor.

#### Phase 1: Clinical Informatics Expert Panel

To establish preliminary validity, six board certified clinical informaticians with experience in AI and clinical decision support (CDS) design, implementation, and evaluation were recruited from the Pediatric CDS Collaborative, a national research collaborative composed of clinical informaticists from nearly 20 pediatric institutions in the US. Each expert rated each survey item assessing how representative and clear the items were with respect to the construct of interest. For example, for the workload construct participants were asked these two question(s):

a. For the overall care of this patient, on a scale of 0 (Low workload)-100 (High Workload) please rate your over all workload Please rate how representative the above question is to understand the workload. (The subjective demands of a particular task).

i. Item is not representative
ii. Item needs major revisions to be representative
iii. Item needs minor revision to be representative
iv. Item is representative
b. For the overall care of this patient, on a scale of 0 (Low workload)-100 (High Workload) please rate your overall workload Please rate how representative the above question is to understand the workload. (The subjective demands of a particular task).

i. Item is not clear
ii. Item needs major revisions to be clear
iii. Item needs minor revision to be clear
iv. Item is clear

Panelists also provided qualitative feedback on redundancy, the clinical interpretability of the instrument items, and the potential for adapting the instrument to diverse CDS interventions.

#### Phase 2: Human Factors Expert Panel

Seven experts with formal training in human factors who have studied human performance related to healthcare were recruited nationally via e-mail. Survey items revised after Phase 1 were first rated individually by these experts using the same type of survey question used in Phase 1 for clarity and representativeness/alignment with established human performance constructs. The panel then participated in four facilitated 90-minute focus group discussions (two for each round) addressing: (1) the scope of the tool (e.g., distinguishing between specific alerts vs broader AI “findings” and “recommendations”), (2) the adequacy and discriminant validity of constructs such as agreement, interpretability, trust, workload, sociotechnical relationships, and unintended consequences, and (3) item wording specifically designed to mitigate cognitive biases, scaling, and response-option structure. After each round, the updated instrument was re-rated for clarity and feasibility until consensus was reached.

### Producing the SHAPE-AI Survey

The final step of the development process involved a thematic synthesis of the qualitative feedback and quantitative ratings collected during the two expert validation phases. Discrepancies between the clinical informatics requirements for operational feasibility (Phase 1) and the human factors requirements for theoretical rigor (Phase 2) were resolved through iterative review by the core research team (authors MVM, MO, SK). Final decisions were made to ensure the instrument could capture the nuance of human-AI teaming across diverse clinical AI applications

### Ethics

This research complied with the American Psychological Association Code of Ethics and was approved by the Institutional Review Board at Children’s Healthcare of Atlanta STUDY00001383

## Results

### Identification of Human Performance Constructs

The initial literature review identified seven core human performance constructs potentially impacted by AI in healthcare: situational awareness, explainability, trust, automation bias, human-computer agreement, workload, and the impact on staff relationships (Table 1). Based on our previously published work, including the initial survey review of human performance constructs and subsequent semi-structured interviews,^21^ we expanded this list to include differential diagnosis, treatment planning, team communication, tool satisfaction, and clinician response behavior. These constructs formed the basis of the initial 10-item survey instrument subjected to expert validation.

**Table 1:** Initial Constructs and Survey Items.

| Construct(s) | Definition | Initial survey items |
| --- | --- | --- |
| Situation Awareness / Impact | Dynamic understanding of ongoing situation through the lens of perception, comprehension and projection.<sup>8</sup> | Did the alert lead to (multiselect)<br>a. more timely care for the patient<br>b. change in your diagnosis or differential<br>c. change in treatment for the patient<br>d. change in staffing or level of care for this patient<br>e. improvement in team communication<br>f. situational awareness |
| Impact | Changes in diagnosis, efficiency, team communication, or confidence |  |
| Workload | Subjective demands of a particular task.<sup>23</sup> | For the overall care of this patient, on a scale of 0 (Low workload)-100 (High Workload) please rate your overall workload |
| Explainability | Characteristics of AI that enable users to know logic | a. How well do you understand why this alert is appearing at the time when it appeared? |
|  | behind decisions and parameters used to make the decisions.<sup>23</sup> | Not at all<br>b. Slightly<br>c. Somewhat<br>d. Mostly<br>e. Completely |
| Human-Computer Agreement | User agreement with the AI system<sup>24</sup> | How surprised are you that this alert showed for the patient?<br>a. Not at all<br>b. Slightly<br>c. Somewhat<br>d. Mostly<br>e. Completely<br>Do you agree with alert that this patient was at high risk for <clinical condition>?<br>a. Not at all<br>b. Slightly<br>c. Somewhat<br>d. Mostly<br>e. Completely<br>When the alert appeared for this patient, how appropriate was the <alert> ?<br>a. Not at all<br>b. Slightly<br>c. Somewhat<br>d. Mostly<br>e. Completely |
| Automation Bias | Tendency to overly rely on automation leading to errors of commission or omission<sup>25,26</sup> | N/A (unable to measure directly from clinician responses) |
| Trust | Trust is the attitude that an AI system will help achieve an individual's goals in a situation characterized by uncertainty and vulnerability.<sup>9</sup><br>Overtrust: Trust exceeds system capabilities, leading to misuse.<sup>9,10</sup><br>Distrust: Trust falls short of system capabilities, leading to disuse.<sup>9,10</sup> | In general, how much do you trust this alert ?<br>a. Not at all<br>b. Slightly<br>c. Somewhat<br>d. Mostly<br>e. Completely |
| Relationship between staff and patients | The way in which people regard and behave toward each other. | How adversely did the alert/huddle impact your relationship with the patient?<br>a. Not at all<br>b. Slightly<br>c. Somewhat<br>d. Mostly<br>e. Completely |
| Satisfaction | The overall contentment, engagement, experience, and happiness. | In general, how satisfied are you with the alert?<br>a. Not at all<br>b. Slightly<br>c. Somewhat<br>d. Mostly<br>e. Completely |
| Alert Performance | The effectiveness and efficiency in delivering relevant, timely, and actionable information to the right clinician through the right channel to improve patient outcomes and safety, without causing undue burden or "alert fatigue | How often do you ignore this alert because of how frequently it is incorrect<br>a. Almost never<br>b. Once in a while<br>c. Sometimes<br>d. Often<br>e. Almost always |

### Phase 1: Clinical Informatics Expert Panel

The Clinical Informatics expert panel evaluated the initial instrument (Table 1) for content validity and clarity. The Clinical Informatics panel rated most items > 0.8 on the CVI, except for four constructs – Impact (CVI = 1.0 [Representativeness]; 0.7 [Clarity]), Human-Computer Agreement (CVI = 0.6 [Representativeness]; 0.8 [Clarity]), Workload (CVI = 0.7 [Representativeness]; 0.7 [Clarity]), and Patient-Staff Relationships (CVI = 0.5 [Representativeness]; 0.7 [Clarity]). Panel feedback highlighted three primary concerns:

- Redundancy in Agreement: Experts noted that the two separate items measuring “human-computer agreement” were conceptually indistinct and recommended merging them into a single assessment of agreement.
- Ambiguity in Workload Measurement: Experts questioned the validity of a single-item workload measure. They argued that respondents may struggle to distinguish the incremental cognitive load of the AI tool (e.g., interpretation and decision-making) from their baseline clinical workload (e.g., clinical complexity).
- Generalizability: The panel recommended replacing AI-specific terminology with broader CDS language to ensure the instrument could evaluate diverse algorithmic interventions (e.g., non-AI rules-based alerts).

### Phase 2: Human Factors Expert Panel

The Human Factors expert panel reviewed the revised instrument Feedback centered on refining construct validity, managing potential cognitive bias, optimizing measurement design, and clarifying the operational scope.

#### Construct Refinement and Measurement Challenges

A primary outcome of Phase 2 was the intentional decoupling of core constructs to mitigate ambiguity and improve discriminant validity, especially for near-real-time assessment.

Experts universally agreed with breaking “Human-Computer Agreement” into two distinct components, helping to identify cases where a clinician agrees with the AI’s finding or risk assessment but disagrees with its suggested action.

1. **Agreement with the AI’s Finding or Risk Assessment:** Evaluating the underlying model’s identification of risk or condition
2. **Agreement with the AI’s Recommendation or Suggested Action:** Evaluating the proposed clinical course

The panel noted that questions about understanding the AI rationale often confuse the technical feature of explainability (the system interface stating how it works) with the cognitive outcome of interpretability (the user’s ability to make sense of the output).

Experts cautioned that relying on self-reported understanding risked measuring the persuasiveness of the interface (sometimes referred to as “storytelling”). Given that perceived transparency is highly variable and can potentially affect clinician interaction with the AI system, the need for subjective assessment via the survey was noted as a valuable leading indicator.

To address the ambiguity identified in Phase 1, the panel recommended a simplified scaling approach focused on the specific outcome of the interaction. Rather than attempting to measure absolute cognitive load, the final question was refined to capture the relative change in burden: whether the AI system created more work, reduced work, or resulted in no additional work.

Discussion highlighted that teaming (which includes communication, coordination, collaboration, and trust) is complex and context-dependent, often requiring specialized methodologies like observation to measure accurately. Panelists agreed that assessing the direct impact on human-to-human team communication and coordination was necessary, particularly in scenarios where the AI agent might intervene or replace existing communication channels. This resulted in items focused on the AI’s influence on provider-patient and provider-team interactions

### Cognitive Biases and Item Formatting

The survey instrument was iteratively modified to address potential respondent bias inherent in self-reporting.

Experts noted that items requiring clinicians to report their cognitive state (e.g., degree of surprise or initial suspicion) were susceptible to recall and confirmation (“saving face”) bias, wherein a respondent might retrospectively alter their pre-decision state to align with the correct AI outcome. To mitigate this, items were reframed to evaluate system performance (e.g., “How clearly did the tool indicate…”) rather than implying a potential failure in user judgment.

For concepts like trust, a graded continuum was recommended over binary (Yes/No) options. Furthermore, panelists noted difficulty in differentiating subtle differences, such as “slightly” versus “somewhat,” on some scales. For scales related to influence or impact, the inclusion of a neutral option, such as “No Effect,” placed centrally between positive and negative poles, was recommended to avoid forcing a response where the AI was not perceived as having an impact.

To ensure the instrument’s longevity, the panel advocated for broadening the terminology beyond “alert” to accommodate diverse CDS modalities, including passive alerts, predictive risk scores, and other AI interventions that offer findings or recommendations. Operational constraints (i.e. interruption to patient care and challenges in administering surveys immediately after AI system interactions) required the survey to be deployed within 24 hours of exposure to the AI tool. Questions focus on a specific patient interaction to minimize recall bias.

### Final Instrument Domains

Following this iterative process, the expert panel reached a final consensus on the SHAPE-AI instrument. The validated domains and a summary of the survey items are presented in Table 2, while the complete survey instrument is provided as a standalone resource in the Appendix 1.

**Table 2:** Final Constructs and Survey Items.

| Construct(s) | Description | Final survey items |
| --- | --- | --- |
| Perceived Impact | Changes in diagnosis, efficiency, team communication, or confidence | 1. Did the AI have an impact on any of the following (multiselect)<br>a. Situational awareness (e.g. seeing new information and/or new understanding of patient risk, anticipate actions you need to take)<br>b. Timely care for the patient<br>c. Patient diagnosis or differential<br>d. Treatment for the patient<br>e. Staffing or level of care for this patient<br>f. Team communication and/or co-ordination<br>g. Confidence in your clinical decisions<br>h. None of the above<br>Free Text Optional |
| Interpretability | Cognitive clarity of the output rationale | 2. How clearly did the AI system explain the reason for its finding? (Interpretability)<br>a. Very Clearly<br>b. Somewhat Clearly<br>c. Not Clearly at All |
|  |  | Free Text Optional |
| Human-Computer Agreement | Assessment of the finding vs. the recommendation. | **3. Did you agree with the AI's finding [risk assessment]? (Agreement)**<br>a. Yes<br>b. No<br>**Free Text (optional)**<br>**4. (If "No" in question 3) Which of the following best explains why you did not fully agree with the AI's finding? (Select all that apply)**<br>a. The AI's finding was based on inaccurate or outdated information.<br>b. The AI's finding did not account for important clinical context or patient factors.<br>c. The AI's findings did not match my own clinical reasoning.<br>d. Other (please specify)<br>**5. Did you agree with the AI's recommendation?**<br>a. Yes<br>b. No<br>**Free Text (optional)**<br>**6. (If "No" to question 5) Please explain why you didn't agree with the AI recommendation.**<br>a. Evidence to the recommendation is weak<br>b. There are other equally good alternative actions<br>c. The action recommendation doesn't meet patient condition.<br>d. The action was not feasible to perform in this clinical setting.<br>e. The action was not specific enough to be useful.<br>f. Other (Please specify) |
| Workload | Relative change in clinical burden. | **7. How did this AI system affect your work? (Workload)**<br>a. Created more work<br>b. Reduced my work<br>c. No additional work |
| Trust | Sentiment toward specific recommendations. | **8. To what extent did you trust this <AI system>? (Trust)**<br>a. Fully trusted<br>b. Somewhat trusted<br>c. Neutral/Unsure<br>d. Did not trust |
| Unexpected outcomes | Open-ended capture of leading indicators for safety, both positive and negative. | **9. Did or could this <AI System> lead to potential unexpected outcomes (reduced cognitive burden, unintended clinician impact or patient safety events/harm, patient/family interactions, health equity etc.)? Multi select**<br>a. Positive Unexpected Outcome(s)<br>b. Negative Unexpected Outcome(s) |
|  |  | c. No Unexpected Outcome<br>Optional description for each option<br>**Free Text (optional)** |
| Relationship between staff and patients | Influence on provider-patient and provider-team interactions. | 10. Compared to baseline, how did the AI system affect your interaction with the patient/family?<br>a. Positively<br>b. Negatively<br>c. No effect at all<br>**Free Text (optional)** |
| Automation Bias | Tendency to overly rely on automation leading to errors of commission or omission<sup>25,26</sup> | Optional:<br>11. Did you verify the AI's finding with other clinical resource (e.g., labs, vitals) or another clinician or your own training/knowledge before acting? (Automation Bias)<br>a. Yes<br>b. No<br>c. Not necessary |

## DISCUSSION

This study developed and iteratively refined a concise survey tool to characterize the near-real-time impact of AI on human performance in clinical settings, explicitly grounding the work in human factors and sociotechnical systems framework. By starting from constructs identified in the literature^18^ and then subjecting a 10-item tool to review by clinical informatics and human factors experts, the work moves beyond ad hoc usability questions toward a more systematic representation of how AI systems shape situational awareness, decision-making, workload, and team relationships. The final domains – perceived impact, agreement with findings and recommendations, interpretability, trust, workload, sociotechnical relationships, and unintended consequences – align with the “Joint Cognitive System” (JCS) framework for human-AI teaming in healthcare, which emphasize that performance is an emergent property of the human-AI partnership rather than the technical output of the model alone.^27,28^

A key contribution of this work is the explicit separation of agreement with the AI’s finding from agreement with its recommended action. Prior studies of AI-enabled decision support often treat acceptance or override as a single behavioral endpoint, obscuring situations where clinicians endorse the risk assessment but deliberately choose a different course of action based on patient preference, comorbidities, or resource constraints.^29,30^ By decoupling these constructs, the instrument serves as a diagnostic tool for identifying specific breakdowns in the human-AI control loop, distinguishing between model miscalibration (poor findings) and clinical misalignment (poor recommendations). Similarly, distinguishing interpretability (clarity of the output rationale in context) from technical explainability acknowledges that human performance depends not on the existence of a rationale, but on the clinician’s ability to apply that rationale to safe and efficient decision-making at the point of care.^13,31^

At the same time, expert feedback highlighted the inherent difficulty of measuring higher-order cognitive constructs such as trust, explainability, and automation bias via brief self-report immediately after a clinical encounter. Consistent with recent reviews, the panels emphasized that trust in AI is dynamic and multidimensional;^32^ perceived trust captured on a rating scale may diverge substantially from “calibrated trust,” in which clinicians align their reliance on the tool with its true performance characteristics. While we do not directly measure calibrated trust, this can be approximated by linking participant response Item 8 with participant alignment in the AI system recommendations and with clinical outcomes. For example, if the model positive predictive value is only 10% but clinicians follow through with treatment decisions for 50% of AI recommendations there is overtrust. Likewise, post-hoc explanations can increase user confidence even when they are driven by spurious features, meaning that self-reported “understanding” may sometimes reflect interface persuasiveness rather than genuine insight into model behavior.^31,33,34^ In response, the final version of the instrument focuses on user sentiment (e.g., perceived interpretability and trust) as leading indicators and deliberately avoids over-claiming them as direct measures of safety or error detection, reinforcing the need to triangulate survey responses with log data, model performance, and outcome measures.

The panels also surfaced important design trade-offs between brevity and construct coverage. To make the instrument deployable within 24 hours of an AI interaction and feasible for routine use, several potentially important constructs – such as detailed error identification and recovery behaviors, training adequacy, and equity impacts – could not be fully elaborated. Instead, they are partially captured through broader items on unintended consequences and free-text responses, which can be used to trigger deeper qualitative inquiry where needed. This design choice aligns with a growing recognition that frontline clinicians are already experiencing substantial cognitive and documentation burden,^35^ and that any additional measurement must be tightly scoped to avoid exacerbating the very workload and alert fatigue issues it aims to study.^36^ The resulting instrument is therefore best understood as a rapid, situational probe that can be layered with observational studies, think-aloud protocols, or ethnographic methods when organizations need richer insight into human-AI teaming.

Several limitations suggest directions for future work. First, the current validation relied on expert panels rather than large-scale psychometric evaluation with end users, so further testing is needed to assess reliability, sensitivity to change, and construct validity across diverse AI tools, use cases, clinical roles, and settings. Second, the instrument was designed primarily around predictive AI and CDS-style interventions; although terminology was broadened to encompass a wider range of AI modalities, additional adaptation will likely be required for generative systems that support documentation, summarization, or patient communication. Third, because the survey captures subjective, scenario-specific perceptions, it cannot on its own quantify calibrated trust, automation bias, or safety outcomes;^34^ these require linkage to event logs, override patterns, error reports, and patient-level outcomes. Finally, while the tool attends to team communication and patient–family relationships, more work is needed to integrate it with complementary methods that capture team cognition and organizational context. Addressing these gaps will be essential to realizing the full potential of the instrument as part of a multi-method toolkit for evaluating and improving human performance with AI in healthcare.

## CONCLUSION

SHAPE-AI represents a significant step toward the systematic and proactive monitoring of human-AI teaming in clinical practice. This concise, expert-validated 10-item instrument is designed to measure the human factors impact of clinical AI in near-real-time. By prioritizing brevity and clarity, the tool is operationally viable for deployment within the high-stakes environment of clinical workflows without inducing survey fatigue. While limitations in self-reporting trust and higher-order cognitive processes remain inherent to survey methodologies, this instrument provides a standardized sociotechnical foundation for assessing how AI interventions shape clinician judgment, workload, and team dynamics. Ultimately, SHAPE-AI allows health systems to detect and mitigate latent failures in their human-AI joint cognitive system. Future work should focus on pairing this subjective measure with technical logs, clinician performance metrics, and patient outcomes to build a holistic and multimodal view of safety, utility, and human-AI collaboration in healthcare.

### Key points

- SHAPE-AI is a brief, 10-item survey designed to capture clinicians’ near-real-time experience of AI-enabled clinical decision support at the point of care.
- The instrument was developed through a construct-driven process integrating literature review, prior qualitative work with sepsis AI users, and sequential input from clinical informatics and human factors experts.
- SHAPE-AI measures key human factors domains: perceived impact, interpretability, agreement with findings, agreement with recommendations, trust, workload, provider–patient and provider–team relationships, unexpected outcomes, and optional automation bias.
- SHAPE-AI is intended to complement technical performance metrics and outcome data, offering health systems a standardized, low-burden tool to detect workflow misalignment, mis-calibrated reliance, and unintended consequences of AI in clinical practice.

## Supporting information

while the complete survey instrument is provided as a standalone resource in the Appendix 1

## Data Availability

Not Applicable

## Notes

**Funding:** This work was supported by the Agency for Healthcare Research and Quality (AHRQ) and the National Center for Advancing Translational Sciences (NCATS) of the National Institutes of Health (NIH) grant numbers 5R03HS029417-02, UL1TR002378, and KL2TR002381.

### Competing Interest Statement

The authors have declared no competing interest.

### Funding Statement

This work was supported by the Agency for Healthcare Research and Quality (AHRQ) and the National Center for Advancing Translational Sciences (NCATS) of the National Institutes of Health (NIH) grant numbers 5R03HS029417-02, UL1TR002378, and KL2TR002381.

### Author Declarations

This study was approved by CHOA institutional review borad STUDY00001383

### Summary of Updates

The complete survey instrument is provided as a standalone resource in the Appendix 1.

