## Supplementary material for "SHAPE-AI: Development and Expert Validation of a Survey for Human–AI Performance Evaluation in Healthcare": while the complete survey instrument is provided as a standalone resource in the Appendix 1

Final Survey Instrument:**

This survey evaluates the AI system's performance, not your clinical skills. Please rate the AI system that you very recently interacted critically on the following:

1. Did the AI have an impact on any of the following (multiselect)
a. Situational awareness (e.g. seeing new information and/or new understanding of patient risk, anticipate actions you need to take)
b. Timely care for the patient
c. Patient diagnosis or differential
d. Treatment for the patient
e. Staffing or level of care for this patient
f. Team communication and/or co-ordination
g. Confidence in your clinical decisions
h. None of the above

**Free Text (optional)**

2. How clearly did the AI system explain the reason for its finding? (Explainability)
a. Very Clearly
b. Somewhat Clearly
c. Not Clearly at All

**Free Text (optional)**

3. **Did you agree with the AI's finding [risk assessment]? (Agreement)**
a. Yes
b. No

**Free Text (optional)**

4. **(If "No" in question 3)** **Which of the following best explains why you did not fully agree with the AI's finding? (Select all that apply)**
a. The AI's finding was based on inaccurate or outdated information.
b. The AI's finding did not account for important clinical context or patient factors.
c. The AI's findings did not match my own clinical reasoning.
d. Other (please specify)
5. **Did you agree with the AI's recommendation?**
a. Yes
b. No

**Free Text (optional)**

6. (If “No” to question 5) Please explain why you didn’t agree with the AI recommendation.
a. Evidence to the recommendation is weak
b. There are other equally good alternative actions
c. The action recommendation doesn’t meet patient condition.
d. The action was not feasible to perform in this clinical setting.
e. The action was not specific enough to be useful.
f. Other (Please specify)
7. How did this AI system affect your work? (Workload)
a. Created more work
b. Reduced my work
c. No additional work

**Free Text (optional)**

8. To what extent did you trust this <AI system>? (Trust)
a. Fully trusted
b. Somewhat trusted
c. Neutral/Unsure
d. Did not trust

**Free Text (optional)**

9. Did or could this <AI System> lead to potential unexpected outcomes (reduced cognitive burden, unintended clinician impact or patient safety events/harm, patient/family interactions, health equity etc.)? Multi select
a. Positive Unexpected Outcome(s)
b. Negative Unexpected Outcome(s)
c. No Unexpected Outcome

Optional description for each option

**Free Text (optional)**

10. Compared to baseline, how did the AI system affect your interaction with the patient/family?
a. Positively
b. Negatively
c. No effect at all
11. Comments / Feedback (Optional)

_______

Additional optional questions

- Did you verify the AI's finding with other clinical resource or another clinician (e.g., labs, vitals) or your own training/knowledge before acting? (Automation Bias)
  a. Yes
  b. No
  c. Not necessary
